# Percutaneous Coronary Intervention vs Coronary Artery Bypass Graft In- hospital Outcomes in Diabetes

**DOI:** 10.1101/2023.03.29.23287929

**Authors:** Qianyun Luo, Renxi Li

## Abstract

**Background:** Despite extensive research on coronary artery bypass surgery (CABG) and percutaneous coronary intervention (PCI) outcome differences, current literature lacks representation of short-term in-hospital outcomes in patients with existing medical conditions. This study aimed to compare perioperative outcomes of these two revascularization procedures in diabetes mellitus (DM) patients.

**Methods:** The National Inpatient Sample (NIS) was used to extract patients who received CABG or PCI surgery between the last quarter of 2015 to 2020 based on ICD10-PCS. Patients of age<40 were excluded for congenital heart defects. Preoperative differences were noted and adjusted using multivariable logistic regression. Adjusted odds ratios (aOR) with 95% confidence intervals (CI) were estimated between PCI and CAGB groups.

**Results:** A total number of 90,662 CABG and 173,725 PCI cases of patients with DM were identified in NIS. Compared to CABG, patients who underwent PCI had increased mortality (2.75% vs 2.00%, aOR 1.266, p<0.0001), myocardial infarction (1.5% vs 1.17%, aOR 1.223, p<0.0001), and were less likely to experience respiratory events (0.38% vs 6.33%, aOR 0.055, p<0.0001), stroke (0.03% vs 0.06%, aOR 0.385, p<0.0001), acute kidney injury (20.37% vs 25.37%, aOR 0.727, p<0.0001), sepsis (0.01% vs 0.05%, aOR 0.172, p<0.0001), shock (0.11% vs 0.74%, aOR 0.139, p<0.0001).

**Conclusions:** The NIS database collects enormous records from nationwide providers, offering great statistical power. PCI was associated with a markedly higher in-hospital mortality rate but a lower morbidity rate in patients with DM as compared to CABG. Therefore, physicians should weigh both mortality and morbidity when considering surgical treatment to DM patients.

## Introduction

Coronary artery disease (CAD) is the leading cause of mortality in the United States, accounting for approximately 610,000 deaths annually.^1^ For CAD patients with an indication for revascularization, two established modalities, coronary artery bypass grafting (CAGB) and percutaneous coronary intervention (PCI), are the two most common surgical procedures employed for myocardial revascularization.^2^ While there is a plethora of studies comparing the outcomes of CABG and PCI, the debate on which procedure provided better results for the treatment of CAD still remains.^3*,4^

Diabetes Mellitus (DM) is a common medical condition affecting approximately 11.3% of the United States population.^1^ CAD is the main cause of death and a major determinant of long-term prognosis for patients with DM.^5^ Patients with DM have a high mortality rate after myocardial infarction (MI) and are at a higher risk of developing CAD with accelerated atherosclerosis and worsened coronary artery stenosis due to factors such as hyperglycemia, lipoprotein abnormalities, and inflammation.^6^ Approximately 25% of CABG and 33% of PCI procedures were performed on patients with DM in the United States, and the results of these procedures are less effective than in patients without DM with a higher risk of restenosis and stent thrombosis.^5^

Previous studies,^3–29^ including large randomized controlled trial Coronary Artery Revascularization in Diabetes (CARDia) trial^11^ and Fibromyalgia Relapse Evaluation and Efficacy for Durability of Meaningful Relief (FREEDOM) trial,^12^ indicated patients with DM who underwent CABG have a lower mortality rate and reduced risk of MI than those who underwent PCI. Despite extensive research on long-term outcome differences of CABG vs PCI in patients with DM, there is very little representation^10^ of short-term in-hospital outcomes, and no studies have been powered to detect in-hospital short-term outcomes between the two revascularization strategies in patients with DM. However, the importance of in-hospital short-term outcomes for clinical decision-making cannot be overstated, as it can inform prompt treatment options and guide physicians to provide the effective in-patient care. Therefore, this study aimed to compare the in-hospital perioperative outcomes of CABG vs PCI in DM patients. This study employed the National (Nationwide) Inpatient Sample (NIS) dataset, which approximates 20% of all discharges from U.S. community hospitals, providing considerable statistical power and allowing for comprehensive analysis of trends and outcomes. Therefore, this study was informative for clinicians to assess and initiate perioperative management accordingly.

## Patients and Methods

### Data Source

This population-based, retrospective observational study utilized data from the US NIS database, which is the largest all-payer, continuous inpatient care database in the United States, obtaining data from about 1,050 hospitals across 44 States in the US, representing a 20% stratified sample of all discharges from U.S. community hospitals as defined by the American Hospital Association.

### Study Population

Patients with DM were identified by Elixhauser Comorbidity^33^ and those underwent CABG or PCI procedures were identified in the database from the last quarter of 2015 to 2020. Patients of age less than 40 were excluded for congenital heart defects. Patients who underwent CABG and PCI were identified using *Internal Classification of Disease, Tenth Revision, Procedural Coding System* (ICD-10-PCS) codes: 0210xxxx for CABG; 02703xx, 02713xx, 02723xx, and 02733xx (027×3Tx and 027×3Zx excluded) for PCI.

### Preoperative Values

Patient demographics and comorbidities were collected and compared between CABG and PCI groups (Table 1). Demographics included race and sex. Comorbidities were identified by Elixhauser Comorbidities^33^, which included AIDS, alcohol use, autoimmune disease, lymphoma, leukemia, metastatic cancer, solid tumor without metastasis in situ, solid tumor without metastasis malignant, cerebral vascular disease, heart failure, dementia, depression, drug abuse, uncomplicated hypertension, complicated hypertension, severe liver disease, chronic lung disease, obese, paralysis, peripheral vascular disease, severe renal failure, hypothyroidism, other thyroid disorders, and valve disease.

**Table 1.** Comorbidities and demographics of patients who underwent CABG or PCI between the last quarter of 2015 and 2020 in NIS database.

|  | **CAGB (n = 90,662)** | **PCI (n = 173,725)** | **p-value** |
| --- | --- | --- | --- |
| **Race, n (%)** |  |  |  |
| White | 64405 (70.55%) | 117884 (66.83%) | <.0001 |
| Black | 7207 (7.89%) | 20053 (11.37%) | <.0001 |
| Hispanic | 9029 (9.89%) | 18939 (10.74%) | <.0001 |
| Asian | 3651 (4%) | 5823 (3.3%) | <.0001 |
| Native Americans | 613 (0.67%) | 1358 (0.77%) | 0.0049 |
| Others | 3044 (3.33%) | 6718 (3.81%) | <.0001 |
| **Sex, n (%)** |  |  |  |
| Male sex | 66221 (72.54%) | 111623 (63.28%) | <.0001 |
| Female sex | 25060 (27.45%) | 64745 (36.71%) | <.0001 |
| **Age, mean ± SD, years old** | **66.18 ± 10.08** | **65.26 ± 12.39** | **<.0001** |
| **Comorbidities, n (%)** |  |  |  |
| AIDS | 292 (0.32%) | 617 (0.35%) | 0.22 |
| Alcohol use | 2179 (2.39%) | 3184 (1.81%) | <.0001 |
| Autoimmune Disease | 2139 (2.34%) | 4292 (2.43%) | 0.1505 |
| Lymphoma | 306 (0.34%) | 725 (0.41%) | 0.0027 |
| Leukemia | 373 (0.41%) | 631 (0.36%) | 0.042 |
| Metastatic cancer | 191 (0.21%) | 887 (0.5%) | <.0001 |
| Solid tumor without metastasis, in situ | 23 (0.03%) | 34 (0.02%) | 0.3298 |
| Solid tumor without metastasis, malignant | 1113 (1.22%) | 2231 (1.26%) | 0.3216 |
| Cerebral vascular disease | 2341 (2.56%) | 4263 (2.42%) | 0.02 |
| Heart Failure | 71 (0.08%) | 69 (0.04%) | <.0001 |
| Dementia | 1051 (1.15%) | 4690 (2.66%) | <.0001 |
| Depression | 8808 (9.65%) | 17455 (9.9%) | 0.0418 |
| Drug abuse | 1354 (1.48%) | 3126 (1.77%) | <.0001 |
| Hypertension complicated | 40847 (44.74%) | 76174 (43.18%) | <.0001 |
| Hypertension uncomplicated | 42917 (47.01%) | 78952 (44.76%) | <.0001 |
| Severe liver disease | 108 (0.12%) | 306 (0.17%) | 0.0005 |
| Chronic lung disease | 19615 (21.49%) | 37112 (21.04%) | 0.0074 |
| Obesity | 33477 (36.67%) | 52150 (29.57%) | <.0001 |
| Paralysis | 1714 (1.88%) | 3349 (1.9%) | 0.7083 |
| Peripheral vascular disease | 11944 (13.08%) | 18315 (10.38%) | <.0001 |
| Severe renal failure | 3472 (3.8%) | 10004 (5.67%) | <.0001 |
| Hypothyroidism | 11138 (12.2%) | 22715 (12.88%) | <.0001 |
| Other thyroid disorders | 1040 (1.14%) | 1536 (0.87%) | <.0001 |
| Valve disease | 866 (0.95%) | 2552 (1.45%) | <.0001 |

### Operative and In-hospital Perioperative Values

Postoperative values considered in this study included in-hospital mortality, stroke, myocardial infarction, major adverse cardiovascular events (MACE), respiratory events, pulmonary embolism, venous thromboembolism, renal failure, acute kidney injury, bleeding events, superficial wound, deep wound, sepsis, shock, length of in-hospital stay greater than 7 days, and transfer. In-hospital mortality was recorded in NIS and other perioperative outcomes were identified using *International Classification of Diseases, Tenth Revision, Clinical Modification* (ICD-10-CM) codes.

### Statistical analysis

Fisher’s exact test was used for categorical variables. Two sample t-test was used for continuous variable. Preoperative differences were noted. Perioperative outcomes were examined by univariate analysis (Fisher’s exact test) and then adjusted for preoperative differences using multivariable logistic regression. Preoperative variables with noted difference (p-value < 0.1) were included in the regression. Adjusted odds ratios (aOR) with 95% confidence intervals (CI) were estimated between PCI and CABG groups. All statistical analyses were performed using SAS (version 9.4) with p < 0.05 considered statistically significant.

### Ethics Statement

Given the use of retrospective, de-identified NIS data, this study excepted from IRB approval by The George Washington University.

## Results

There were 90,662 patients with DM who underwent CABG and 173,725 who underwent PCI identified in NIS between the last quarter of 2015 and 2020. The comorbidities and demographics of the patients were summarized in Table 1. Compared to CABG, there were more female (36.71% vs 27.45%, p < 0.0001), Black (11.37% vs 7.89%, p < 0.0001), Hispanic (10.74% vs 9.89%, p < 0.0001), and Native Americans (0.77% vs 0.67%, p = 0.0049) in DM patients who underwent PCI (Table 1). In contrast, White and Asian DM patients were more likely to undergo CABG than PCI (70.55% vs 66.83%, p < 0.0001; 4% vs 3.3%, p < 0.0001). CABG patients (66.18 ± 10.08 years old) were older (p < 0.0001) than PCI patients (65.26 ± 12.39 years old).

There were no statistically significant differences in AIDS, autoimmune disease, solid tumor, or paralysis between the two groups. DM patients with alcohol use (1.81% vs 2.39%, p < 0.0001), leukemia (0.36% vs 0.41%, p = 0.042), cerebral vascular disease (2.42% vs 2.56%, p = 0.02), heart failure (0.04% vs 0.08%, p < 0.0001), complicated (43.18% vs 44.74%, p < 0.0001) and uncomplicated (44.76% vs 47.01%, p < 0.0001) hypertension, chronic lung disease (21.04% vs 21.49%, p = 0.0074), obese (29.57% vs 36.67%, p < 0.0001), peripheral vascular disease (10.38% vs 13.08%, p < 0.0001), and other thyroid disorders (0.87% vs 1.14%, p < 0.0001) were less likely to undergo PCI than CABG. On the other hand, patients with the following comorbidities were more likely to undergo PCI than CABG: metastatic cancer (0.5% vs 0.021%, p < 0.0001), lymphoma (0.41% vs 0.34%, p = 0.0027), dementia (2.66% vs 1.15%, p < 0.0001), depression (9.9% vs 9.65%, p = 0.0418), drug abuse (1.77% vs 1.48%, p < 0.0001), severe liver disease (0.17% vs 0.12%, p = 0.0005), severe renal failure (5.67% vs 3.8%, p < 0.0001), hypothyroidism (12.88% vs 12.2%, p < 0.0001), and valve disease (1.45% vs 0.95%, p < 0.0001).

The univariate and multivariable logistic regression comparing the in-hospital perioperative outcomes of CABG and PCI were summarized in Table 2 and Table 3, respectively. Compared to CABG, DM patients who underwent PCI had increased in-hospital mortality (2.75% vs 2.00%, aOR 1.266, p < 0.0001), myocardial infarction (1.5% vs 1.17%, aOR 1.223, p < 0.0001), and MACE (1.53% vs 1.27%, aOR 1.154, p < 0.0001). In addition, DM patients who went under PCI were less likely to experience stroke (0.03% vs 0.06%, aOR 0.385, p < 0.0001), respiratory events (0.38% vs 6.33%, aOR 0.055, p < 0.0001), venous thromboembolism (0.41% 0.66%, aOR 0.605, p < 0.0001), acute kidney injury (20.37% vs 25.37%, aOR 0.727, p < 0.0001), renal failure (0.05% vs 0.81%, aOR 0.068, p < 0.0001), deep wound (0.01% vs 0.2%, aOR 0.055, p < 0.0001), sepsis (0.01% vs 0.05%, aOR 0.172, p < 0.0001), shock (0.11% vs 0.74%, aOR 0.139, p < 0.0001), length of stay greater than 7 days (13.93% vs 58.02%, aOR 0.089, p < 0.0001), or transfer (9.47% vs 24.72%, aOR 0.254, p < 0.0001). No difference was observed in perioperative pulmonary embolism, bleeding events, or superficial wounds.

**Table 2.** Univariate analysis of peri-operative outcomes of patients who underwent CABG or PCI between the last quarter of 2015 and 2020 in NIS database.

| Outcomes, n (%) | CAGB (n = 90,662) | PCI (n = 173,725) | p-value |
| --- | --- | --- | --- |
| Mortality | 1828 (2%) | 4842 (2.75%) | <.0001 |
| Stroke | 59 (0.06%) | 47 (0.03%) | <.0001 |
| Myocardial infarction | 1072 (1.17%) | 2649 (1.5%) | <.0001 |
| MACE | 1155 (1.27%) | 2702 (1.53%) | <.0001 |
| Respiratory events | 5775 (6.33%) | 664 (0.38%) | <.0001 |
| Pulmonary embolism | 26 (0.03%) | 60 (0.03%) | 0.496 |
| Venous thromboembolism | 606 (0.66%) | 728 (0.41%) | <.0001 |
| Renal failure | 742 (0.81%) | 95 (0.05%) | <.0001 |
| Acute kidney injury | 23161 (25.37%) | 35926 (20.37%) | <.0001 |
| Bleeding events | 160 (0.18%) | 232 (0.13%) | 0.0055 |
| Superficial wound | 616 (0.67%) | 1160 (0.66%) | 0.6155 |
| Deep wound | 185 (0.2%) | 19 (0.01%) | <.0001 |
| Sepsis | 46 (0.05%) | 16 (0.01%) | <.0001 |
| Shock | 678 (0.74%) | 188 (0.11%) | <.0001 |
| Length of stay >7 days | 52964 (58.02%) | 24569 (13.93%) | <.0001 |
| Transfer | 22565 (24.72%) | 16704 (9.47%) | <.0001 |

**Table 3.** Multivariable logistic regression analysis of peri-operative outcomes of patients who underwent CABG or PCI between the last quarter of 2015 and 2020 in NIS database. Demographics and comorbidities with large difference (p-value < 0.1) were included in the regression.

| Outcomes | aOR for PCI/CABG | Lower 95% CI | Upper 95% CI | p-value |
| --- | --- | --- | --- | --- |
| Mortality | 1.266 | 1.197 | 1.338 | <.0001 |
| Stroke | 0.385 | 0.262 | 0.567 | <.0001 |
| Myocardial infarction | 1.223 | 1.137 | 1.315 | <.0001 |
| MACE | 1.154 | 1.076 | 1.239 | <.0001 |
| Respiratory events | 0.055 | 0.051 | 0.06 | <.0001 |
| Pulmonary embolism | 1.179 | 0.744 | 1.868 | 0.484 |
| Venous thromboembolism | 0.605 | 0.542 | 0.675 | <.0001 |
| Renal failure | 0.068 | 0.055 | 0.085 | <.0001 |
| Acute kidney injury | 0.727 | 0.712 | 0.742 | <.0001 |
| Bleeding events | 0.754 | 0.615 | 0.924 | 0.0065 |
| Superficial wound | 0.923 | 0.836 | 1.02 | 0.1148 |
| Deep wound | 0.055 | 0.034 | 0.088 | <.0001 |
| Sepsis | 0.172 | 0.097 | 0.303 | <.0001 |
| Shock | 0.139 | 0.118 | 0.163 | <.0001 |
| Length of stay >7 days | 0.089 | 0.087 | 0.091 | <.0001 |
| Transfer | 0.254 | 0.248 | 0.261 | <.0001 |

### Comment

Previous studies have compared perioperative outcomes in patients with DM treated with either CABG or PCI;^3–29^ however, only a few focused on short-term in-hospital outcomes^7,8,10,24–29^ with limited data size. The objective of this study was to assess the in-hospital perioperative outcomes of CABG vs PCI in patients with DM to direct clinical decisions for surgery, monitor prognosis, and guide post-surgical management in this patient population. Our results showed minority groups such as female, Black, Hispanic, Native Americans were more likely to undergo PCI over CABG (Table 1). This disparity may be due to PCI having a shorter length of in-hospital stay (Table 2 and 3), and in turns, less financial burden for these patients. Patients with underlying heart, lung, and peripheral vascular diseases were more likely to undergo CABG. In contrast, patients with liver and renal conditions were more likely to undergo PCI surgery; this may be due to concern for compromised peri-operative recovery for the more invasive CABG.

Despite limited research on short-term outcomes, studies that examine in-hospital perioperative outcomes of patients with DM who underwent PCI or CABG have produced conflicting results. Ben-Gal et al reported higher mortality (4.7% vs 1.6%, p = 0.0003) and MI incidence (11.7% vs 7.1%, p = 0.003) in patients who underwent CABG as compared to those who underwent PCI.^24^ However, Zheng et al reported lower mortality (0.3% vs 0.8%, p = 0.03) among the patients who underwent CABG.^28^ Both Zheng et al and Ramanathan et al found a lower rate of MI (2.1% vs 3.1%, p = 0.05; 1.1% vs 4.5 %, p < 0.01) in patients who underwent CABG as compared to those who underwent PCI, ^27,28^ which aligned with our findings (Table 3). Other studies demonstrated no significance in mortality or MI difference between patients who underwent either of the two revascularizations procedures.^8,10,26^ Stroke was also demonstrated as insignificant between the two procedures.^24,26,28^ Despite inconsistency in mortality and MI, our study showed similar findings as Ben-Gal et al who demonstrated higher acute kidney injury (36.1% vs 16%, p < 0.0001) and bleeding events (54.1% vs 9.8%, p < 0.0001) in the CABG group as compared to PCI.^24^ Inconsistency in findings could be attributed to underpowered sample sizes from previous studies.

Our findings indicated while morbidities were lower for patients who underwent PCI compared to CABG, mortality, MI, and MACE incidence were higher (Tables 2 and 3). These findings aligned with the long-term survival benefit of CABG.^10–12^ We hypothesized the leading cause of mortality is cardiovascular-related death such as MI and MACE; however, further research is needed to investigate the underlying cause. While minimally invasive PCI might be chosen to avoid complications compared to CABG, the percentage of complete revascularization is lower,^26^ which leads to a greater risk of cardiac death and potential risk of reoperation for repeated revascularization. Furthermore, sampling bias may contribute to the unobserved morbidities in patients who underwent PCI, since mortality was higher in PCI and deceased patients were excluded from the morbidity comparison. PCI is a less invasive procedure than CABG and may be preferred for patients who are not ideal candidates for complex surgery. As a result, patients who receive PCI may be sicker or have more comorbidities than those who receive CABG. Accelerated vascular aging, arterial stiffening, and arteriosclerosis in DM patients also cannot be discounted as potential explanations for increased mortality of PCI.^30^ These findings had important implications for clinical practice. Clinicians should carefully consider the risks and benefits of each revascularization strategy when making treatment decisions for DM patients. Future research should be conducted to evaluate the primary cause of death, such as by incomplete revascularization or by morbidity, following the procedures of PCI or CABG.

There are a number of limitations of this study. As shown by numerous prior studies, NIS does not capture all pertinent intraoperative parameters – such as the coronary segment directly affected, the diameter of the stenosis, right or left dominance of the coronary arteries, the presence of a lesion, calcification, and diffusion to small vessels – that contribute to successful revascularization and can have a significant impact on morbidity and mortality.^2,19,26,28^ In addition, NIS does not record lab values. Notedly, the hemoglobin A1c level of patients with DM is major aspect that may affect the mortality rate of both revascularization procedures.^31,32^

Overall, the NIS database is a valuable source of information, containing vast amounts of data from providers across the United States. This provides researchers with considerable statistical power, allowing for a comprehensive analysis of trends and outcomes. Our study shows PCI was associated with a markedly higher in-hospital mortality rate but a lower morbidity rate in patients with DM as compared to CABG. Therefore, healthcare providers should take into account both the individual patient’s medical history and overall health status when deciding between CABG and PCI, and providers should consider the potential risks and benefits of each intervention in order to provide the best possible care for patients with DM and CAD.

## Data Availability

All data produced in the present study are available upon reasonable request to the authors

## Acknowledgements

The authors acknowledge the guidance and support of colleagues and reviewers who provided useful feedback throughout the work. The authors also thank our loved ones whose patience and support helped us to complete this research.

